# The effect of balanced energy-protein supplementation provided to lactating women on maternal and infant outcomes: study protocol for a prospectively planned individual patient data (IPD) meta-analysis

**DOI:** 10.1101/2023.11.06.23298006

**Authors:** Mihaela A. Ciulei, Shouhao Zhou, Kelly Gallagher, Sunita Taneja, Nita Bhandari, Patrick Kolsteren, Ameer Muhammad, James M Tielsch, Alemayehu Argaw, Ranadip Chowdhury, Parul Christian, Trenton Dailey-Chwalibóg, Brenda de Kok, Daniel J. Erchick, Fyezah Jehan, Joanne Katz, Subarna Khatry, Carl Lachat, Tsering P Lama, Muhammad Imran Nisar, Yasir Shafiq, Ravi Upadhyay, Alison D Gernand, Maternal BEP Studies Harmonization Initiative

**Author notes:** **Corresponding Author:** Alison D. Gernand, 110 C Chandlee Lab, The Pennsylvania State University, University Park, PA 16802.

## Abstract

**Background:** The high prevalence of infant stunting and maternal undernutrition in low- and middle-income countries poses a significant public health threat. The World Health Organization recommends balanced energy-protein (BEP) supplementation to pregnant women from populations with a high prevalence of underweight (prepregnancy BMI <18.5 kg/m^2^), leaving a notable gap in guidance for lactating women. To address this problem, we established the Maternal BEP Studies Harmonization Initiative (BEP Initiative) to investigate the impact of BEP supplementation given to pregnant and/or lactating women on maternal and infant outcomes by synthesizing data from multiple clinical trials. This is a study protocol for our prospective individual participant data (IPD) meta-analysis on BEP lactation trials.

**Methods:** Data from four randomized controlled trials that include mother-infant dyads in India (n=816), Pakistan (n=957), Burkina Faso (n=800), and Nepal (n=726) will be pooled and analyzed. Women were randomized to BEP (one trial had a third arm with maternal BEP plus infants receiving azithromycin) or control groups at baseline (during the first week) and received the intervention through six months postpartum. A one-stage IPD meta-analysis will be done using mixed-effects linear and log-binomial regression models to account for between-trial heterogeneity. The primary outcome of infant length-for-age z scores (LAZ) at six months of age and secondary outcomes of maternal and infant indicators of nutritional status at six months of age will be examined. Also, we will examine baseline characteristics as covariates and effect modifiers for the BEP to outcome relationship. Risk of bias assessments will be carried out for each of the individual trials using the Cochrane risk of bias tool.

**Discussion:** This prospective IPD meta-analysis uses a one-stage IPD meta-analysis, which allows for higher statistical power to examine outcomes, more flexibility in defining variables, and has the ability to examine many individual- and study-level variables as effect modifiers, allowing conclusions on which individuals or populations may benefit more from BEP given during lactation.

**Trial registration:** This protocol was pre-registered in Open Science Framework (https://osf.io/9nq7z)

## BACKGROUND

Pregnant and lactating women with undernutrition are at high risk of adverse maternal and infant health outcomes. Often, undernutrition is characterized by low body mass index (BMI), low mid-upper arm circumference (MUAC), short stature, and/or micronutrient deficiencies, which put pregnant women at risk for complications such as intrauterine growth restriction and preterm birth (1). The World Health Organization (WHO) recommends balanced energy-protein (BEP) supplementation in populations at risk of underweight (defined as more than 20% of pregnant women with a BMI <18.5 kg/m^2^) to reduce the risk of stillbirth and small for gestational age neonates (2). However, the consequences of undernutrition among lactating women have commonly been overlooked. Lactating women require additional calories to produce milk, and maternal weight status may impact the volume of milk produced and available for infant consumption (3). In food insecure settings, particularly in low- and middle-income settings, there is a high prevalence of infant stunting (4–6). Stunting, or impaired growth due to inadequate nutrition, can have serious long-term consequences for children’s health and development. Meanwhile, breast milk is often the sole or main source of calories and nutrients for infants under six months of age in low-resource settings, and supplementing the diet of women who are breastfeeding could have a direct impact on infant growth and health.

BEP supplementation products are ready-to-use or prepared foods that provide energy and protein (accounting for less or equal to 25% of the total energy content) (7). When given during pregnancy, packaged BEP products are often fortified with multiple micronutrients or if BEP is in the form of locally-prepared food, it is often given along with a multiple micronutrient or iron and folic acid (IFA) tablet. The current evidence in systematic reviews and meta-analyses has focused on studies where BEP supplements are given during pregnancy, and synthesis is lacking for trials giving BEP during lactation (8–12).

To align BEP product formulations, the Bill & Melinda Gates Foundation (BMGF) assembled an expert panel in 2017 to develop guidelines for the macro- and micronutrient content of BEP supplements for pregnancy and lactation (7). The panel also recommended that BEP products be developed and evaluated in both pregnant and lactating women in low-resource settings to assess health benefits. To further advance the evidence-based research, BMGF funded several independent RCTs and in 2020, convened the Maternal Nutrition Harmonization Workshop to harmonize key variables across these trials and prioritize outcomes for the IPD meta-analysis (13). This later led to the formation of the Maternal BEP Studies Harmonization Initiative (hereinafter BEP Initiative) to examine the pooled effect of BEP in pregnancy and lactation on maternal and child health.

The current protocol describes the objectives, data, and analysis plan of our prospective individual participant data (IPD) meta-analysis that focuses on the effect of BEP supplementation given during lactation in four trials with similar designs, outcome measures, and settings (i.e., low-and middle-income countries (LMIC)). The primary aim of this IPD meta-analysis is to assess the effect of BEP supplementation in lactating women on infant length-for-age z scores (LAZ) at six months of age. For secondary outcomes, we will assess maternal and infant weight and malnutrition indicators at six months postpartum. Last, we will examine maternal and infant characteristics that may modify the relationship between BEP intervention and outcomes.

## METHODS/DESIGN

### Protocol and registration

This protocol was preregistered in Open Science Framework (https://osf.io/9nq7z) and all individual trials were prospectively registered online (**Table 1**). We followed The Preferred Reporting Items for Systematic Reviews and Meta-Analysis-Protocol framework, and the checklist is appended.(14) The reporting of the IPD meta-analysis will use the PRISMA-IPD reporting checklist (15).

**Table 1:**
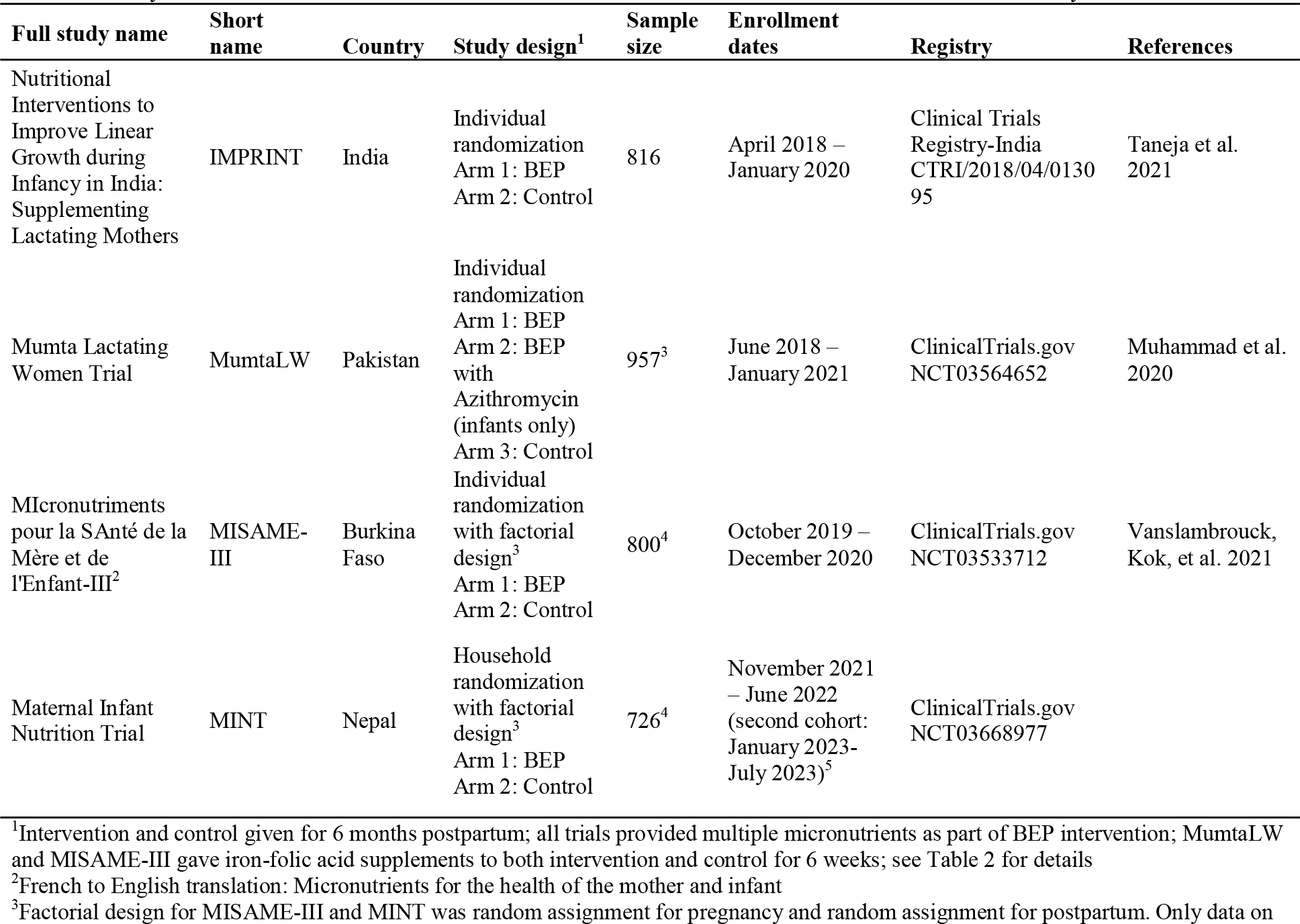
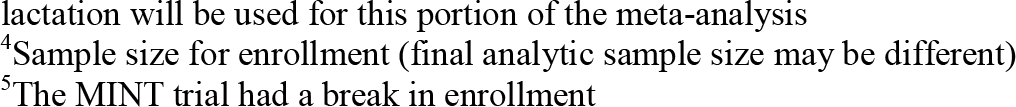
Study information for the maternal BEP lactation trials that will be included in the IPD meta-analysis

| Full study name | Short name | Country | Study design<sup>1</sup> | Sample size | Enrollment dates | Registry | References |
| --- | --- | --- | --- | --- | --- | --- | --- |
| Nutritional Interventions to Improve Linear Growth during Infancy in India: Supplementing Lactating Mothers | IMPRINT | India | Individual randomization<br>Arm 1: BEP<br>Arm 2: Control | 816 | April 2018 – January 2020 | Clinical Trials Registry-India<br>CTRI/2018/04/013095 | Taneja et al. 2021 |
| Mumta Lactating Women Trial | MumtaLW | Pakistan | Individual randomization<br>Arm 1: BEP<br>Arm 2: BEP with Azithromycin (infants only)<br>Arm 3: Control | 957<sup>3</sup> | June 2018 – January 2021 | ClinicalTrials.gov<br>NCT03564652 | Muhammad et al. 2020 |
| Micronutriments pour la SANTé de la Mère et de l'Enfant-III<sup>2</sup> | MISAME-III | Burkina Faso | Individual randomization with factorial design<sup>3</sup><br>Arm 1: BEP<br>Arm 2: Control | 800<sup>4</sup> | October 2019 – December 2020 | ClinicalTrials.gov<br>NCT03533712 | Vanslambrouck, Kok, et al. 2021 |
| Maternal Infant Nutrition Trial | MINT | Nepal | Household randomization with factorial design<sup>3</sup><br>Arm 1: BEP<br>Arm 2: Control | 726<sup>4</sup> | November 2021 – June 2022 (second cohort: January 2023-July 2023)<sup>5</sup> | ClinicalTrials.gov<br>NCT03668977 |  |

<sup>1</sup>Intervention and control given for 6 months postpartum; all trials provided multiple micronutrients as part of BEP intervention; MumtaLW and MISAME-III gave iron-folic acid supplements to both intervention and control for 6 weeks; see Table 2 for details
<sup>2</sup>French to English translation: Micronutrients for the health of the mother and infant
<sup>3</sup>Factorial design for MISAME-III and MINT was random assignment for pregnancy and random assignment for postpartum. Only data on
lactation will be used for this portion of the meta-analysis
<sup>4</sup>Sample size for enrollment (final analytic sample size may be different)
<sup>5</sup>The MINT trial had a break in enrollment

### Trials and sample size

Four BEP supplementation trials during lactation will be included in this prospective IPD meta-analysis located in: India (IMPRINT), Burkina Faso (MISAME-III), Pakistan (MumtaLW), and Nepal (MINT) (see **Table 1** for study information and acronym definitions). All four studies are part of the BEP Initiative. Recruitment is complete for IMPRINT, MumtaLW, and MISAME-III, and sample sizes are as follows: IMPRINT (n=816), MumtaLW (n=957), MISAME-III (n=800), and MINT (n=726). The expected total sample size is 3,299 women. The sample size for some exploratory outcomes will be lower than the total enrollment sample size due to certain information (e.g., blood collection or analysis) being collected on a subset of participants (by design). Also, the sample size for the MISAME-III and MINT trials is lower than the sample size for the full trials because these trials have a factorial design to provide BEP during pregnancy and/or lactation. Groups receiving BEP supplementation during pregnancy are not included in this analysis. Risk of bias assessments will be carried out and reported for each of the individual trials using the Cochrane risk of bias tool (16). There is no risk of duplicate data.

### Power calculations

For this prospective meta-analysis, we assume the mean LAZ at six months of age to be -0.57 (standard deviation 1.10) in the control group(17). A total sample size of 3,299 (1,809 for BEP package group and 1,490 for control group) yields 88% power to detect a minimum LAZ difference of 0.10 between BEP and control at a significance level of 5% using mixed-effects linear regression, and assuming an intra-class correlation as low as 0.30. For secondary outcomes, we will have 89% power to detect a standardized mean difference of 0.10, assuming an intra-class correlation as low as 0.20. The PASS Software v22.0.4 was used for sample size calculation (NCSS, LLC, Kaysville, Utah).

### Participants, eligibility, and study design

In the IMPRINT trial, mother-infant dyads were included in the study if a participant initiated breastfeeding within seven days postpartum. In the MumtaLW trial, lactating women with undernutrition (MUAC <23.0 cm), between 13-49 years of age, and their newborns (captured within seven days from birth) were included in the study. Additionally, lactating women had to intend to exclusively breastfeed the infant for the first six months of age. In the MISAME-III and MINT trials, women between 15-40 and 15-30 years, respectively, were enrolled in the study following a positive urine test if they were found to be missing menstruation in the prior five weeks and following an ultrasound examination that revealed an intrauterine pregnancy <21 completed weeks of gestation. They were in the trial from early gestation through 6 months postpartum and were counseled to exclusively breastfeed for 6 months. Additionally, in all trials, participants indicated that they were not allergic to BEP ingredients (e.g., peanuts).

**Table 1** summarizes the basic study information for included maternal BEP lactation trials. Among them, IMPRINT followed an individual randomization, controlled efficacy trial design. MumtaLW followed a multi-arm, community-based randomized controlled, open-label, assessor-blinded superiority trial design with a treatment allocation ratio of 1:1:1. MISAME-III was an individual randomized 2×2 factorial efficacy design where participants are individually and randomly allocated to a prenatal intervention or control and a postnatal intervention or control group. Similarly, MINT followed a household randomized 2×2 factorial efficacy design where participants in the same household obtained the same prenatal and postnatal intervention or control. The current analyses will focus only on the postpartum intervention.

### Intervention

The intervention tested in each trial was a BEP supplement given to women from birth (or within a week of birth) to six months postpartum. The intervention groups also received what the control group received as the standard of care (see below) in addition to BEP. IMPRINT’s nutritional intervention was five different BEP snacks produced by a local company and a separate multiple micronutrient supplement (**Table 2**) (18). The MumtaLW trial provided a BEP local product called Mumta that is fortified with multiple micronutrients; two sachets per day were provided (19). MumtaLW had a third intervention arm in which the women received BEP and the infants also received one dose of azithromycin at 42 days. The MISAME-III and MINT trials provided a micronutrient fortified BEP supplement produced by the Nutriset (Malaunay, France) (20).

**Table 2:**
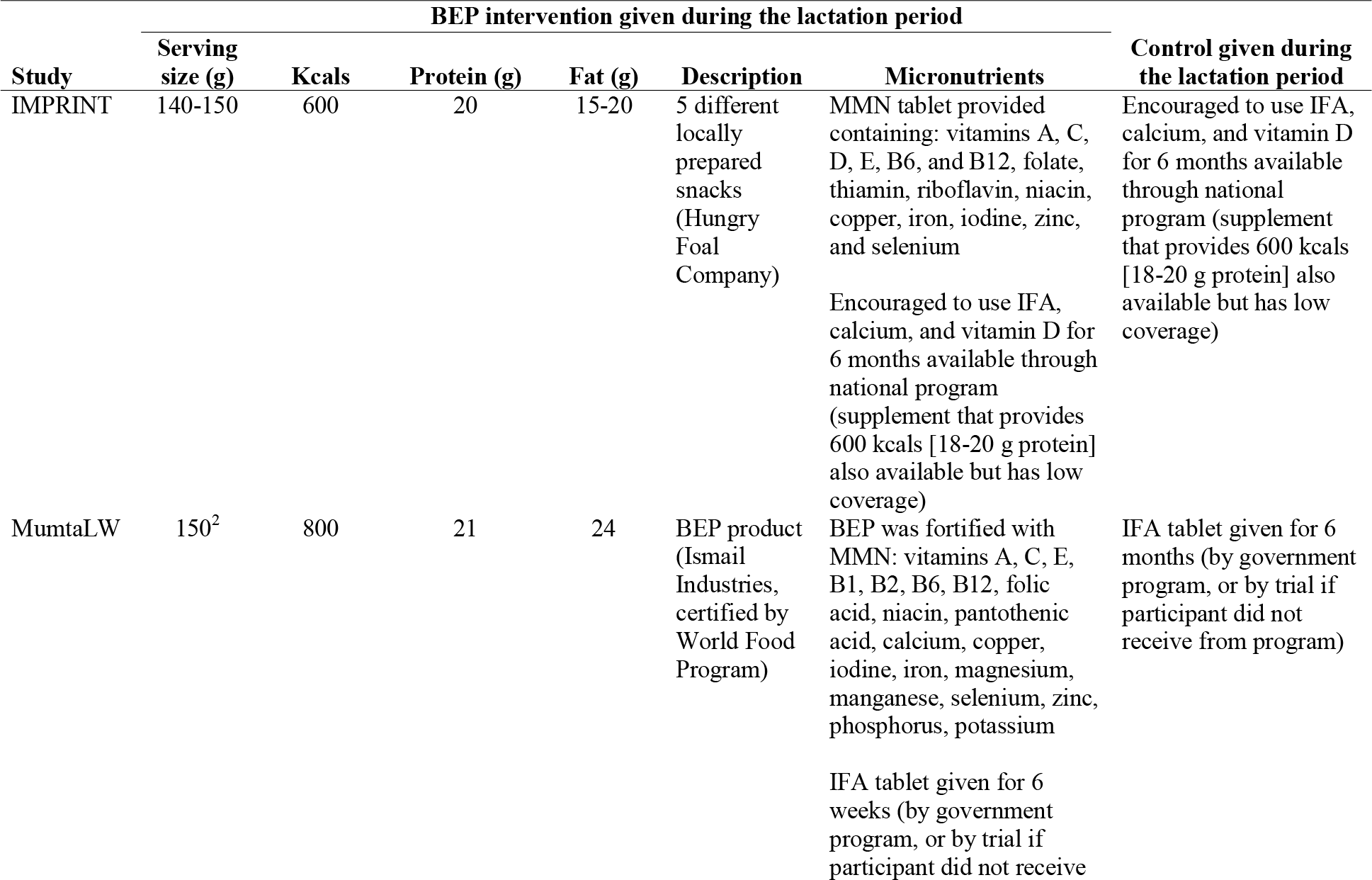
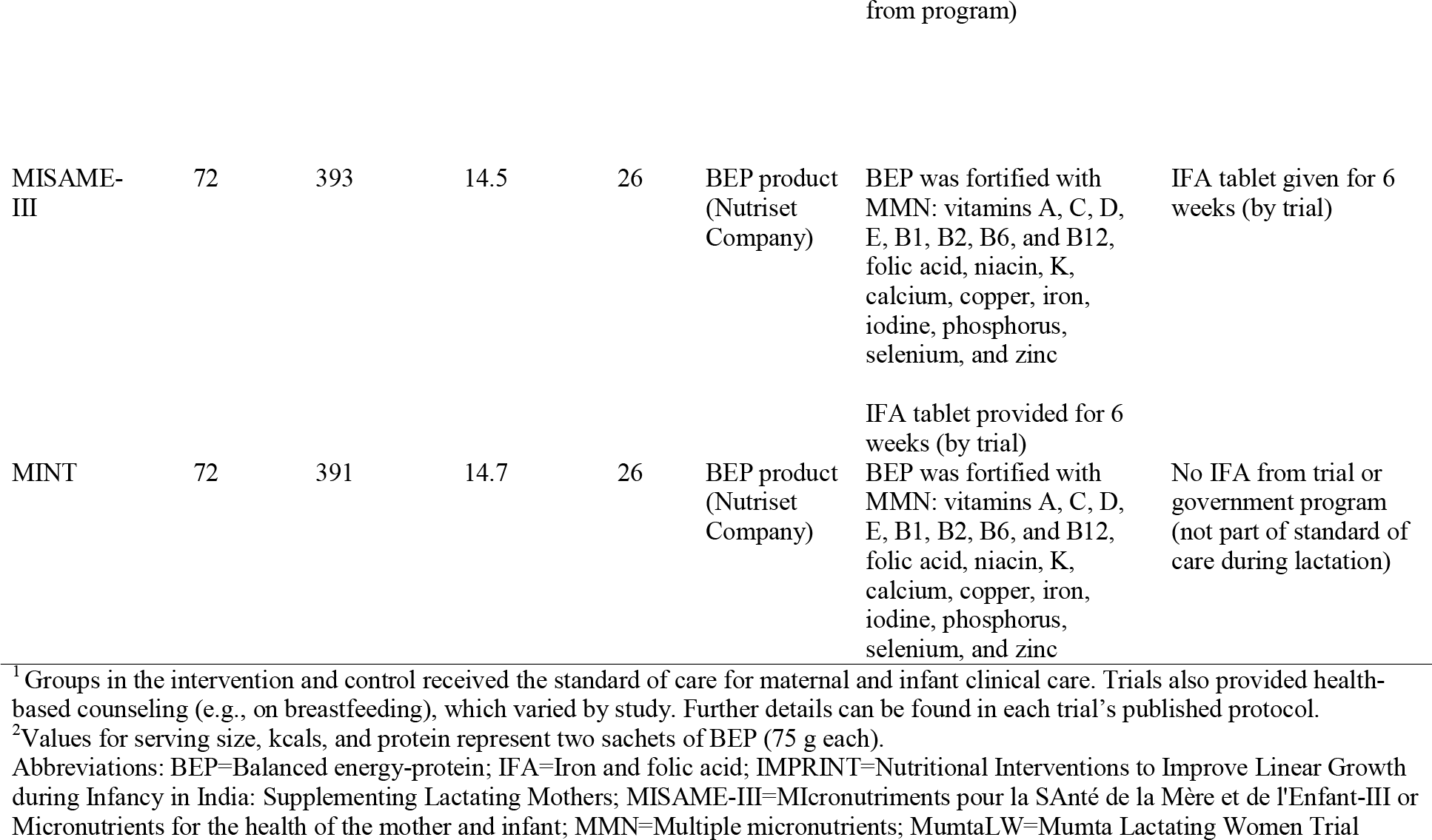
BEP and control trial arm descriptions of energy and nutrients provided to women in the lactation trials that will be included in the IPD meta-analysis^**1**^

### Control/Standard of care

All four trials had a control arm that was intended to align with the standard of care for postpartum women in each country (Table 2). In the IMPRINT trial, women were encouraged to use IFA, calcium, and vitamin D supplements for six months postpartum from the national program in India (i.e., not provided by the trial) (18). The national program also provides a food supplement that contains 600 kcals (18-20 g protein). In the MumtaLW trial, women received IFA from the government program or the trial (i.e., the trial ensured it was provided) for six weeks postpartum. In the MISAME-III trial, women also received IFA from the trial for six weeks postpartum. Finally, in the MINT trial, IFA was not provided as it was not part of the standard of care during lactation in Nepal.

Most trials provided participants (intervention and control) with counseling for nutrition, breastfeeding, and infant care, or referred to services where counseling was available. Trials also encouraged women to use postnatal clinical care for themselves and their infants and referred to clinical services in the case of illness.

### Outcomes and prioritization

The primary outcome for this IPD meta-analysis is infant length-for-age z score (LAZ) at six months of age, which represents linear growth across that timeframe. Secondary outcomes include infant weight and malnutrition at six months of age (weight-for-length z score (WLZ) and weight-for-age z score (WAZ)), infant growth velocity (i.e., change in length and weight), infant stunting (LAZ <-2), wasting (WLZ <-2), and underweight (WAZ <-2)) at six months of age. All infant weight and nutrition status indices (LAZ, WAZ, WLZ) will be calculated based on WHO Growth Standards (21) (or INTERGROWTH-21^st^ for infants born preterm, when gestational age is available), which are sex-specific. We will also assess maternal anthropometry including mean BMI and MUAC, underweight (BMI <18.5 kg/m^2^) and low MUAC (<23.0 cm) at six months postpartum (**Table 3**).

**Table 3:**
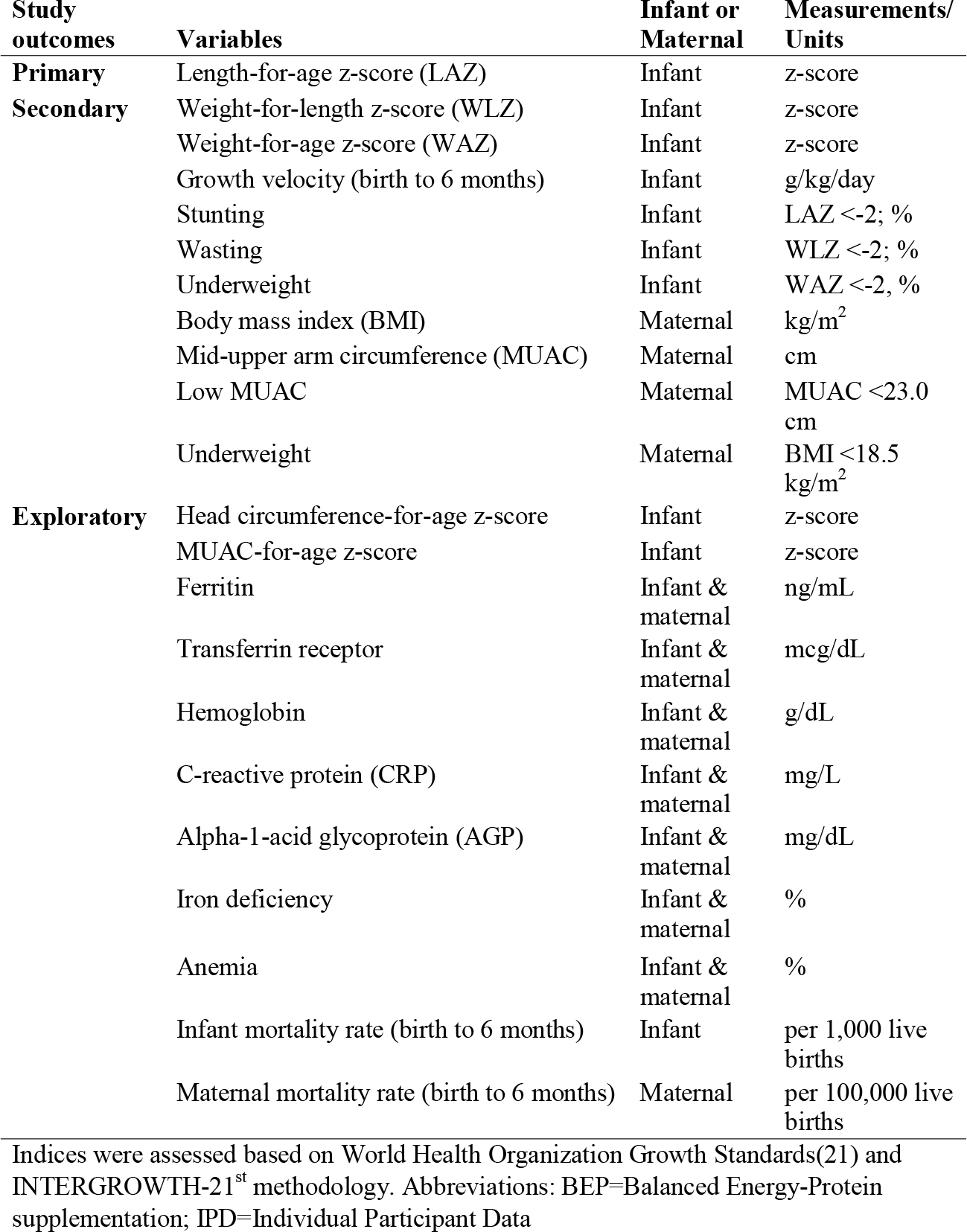
Maternal and infant outcomes (at six months of infant age) to be assessed in the IPD meta-analysis of BEP lactation trials

| **Study outcomes** | **Variables** | **Infant or Maternal** | **Measurements/ Units** |
| --- | --- | --- | --- |
| **Primary** | Length-for-age z-score (LAZ) | Infant | z-score |
| **Secondary** | Weight-for-length z-score (WLZ) | Infant | z-score |
|  | Weight-for-age z-score (WAZ) | Infant | z-score |
|  | Growth velocity (birth to 6 months) | Infant | g/kg/day |
|  | Stunting | Infant | LAZ <-2; % |
|  | Wasting | Infant | WLZ <-2; % |
|  | Underweight | Infant | WAZ <-2, % |
|  | Body mass index (BMI) | Maternal | kg/m<sup>2</sup> |
|  | Mid-upper arm circumference (MUAC) | Maternal | cm |
|  | Low MUAC | Maternal | MUAC <23.0 cm |
|  | Underweight | Maternal | BMI <18.5 kg/m<sup>2</sup> |
| **Exploratory** | Head circumference-for-age z-score | Infant | z-score |
|  | MUAC-for-age z-score | Infant | z-score |
|  | Ferritin | Infant & maternal | ng/mL |
|  | Transferrin receptor | Infant & maternal | mcg/dL |
|  | Hemoglobin | Infant & maternal | g/dL |
|  | C-reactive protein (CRP) | Infant & maternal | mg/L |
|  | Alpha-1-acid glycoprotein (AGP) | Infant & maternal | mg/dL |
|  | Iron deficiency | Infant & maternal | % |
|  | Anemia | Infant & maternal | % |
|  | Infant mortality rate (birth to 6 months) | Infant | per 1,000 live births |
|  | Maternal mortality rate (birth to 6 months) | Maternal | per 100,000 live births |

Indices were assessed based on World Health Organization Growth Standards(21) and INTERGROWTH-21<sup>st</sup> methodology. Abbreviations: BEP=Balanced Energy-Protein supplementation; IPD=Individual Participant Data

Exploratory outcomes include infant head circumference-for-age and MUAC-for-age z scores (based on WHO Growth Standards, see above) at six months; maternal and infant biomarkers for iron deficiency, anemia, inflammation, and other micronutrients at six months of infant age (ferritin, transferrin receptor, hemoglobin, C-reactive protein (CRP), alpha-1-acid glycoprotein (AGP), other micronutrients (biomarkers to be determined)); and maternal mortality rate (per 100,000 live births) and infant mortality rate (per 1,000 live births; Table 3).

### Individual- and study-level covariates

Individual-level variables collected by trials for women include maternal age, education, baseline weight and height, parity, adherence to BEP, health and pregnancy history, diet, breastfeeding type (exclusive, predominant, and partial), and household food insecurity. For infants, variables include sex, gestational age at birth, age at each measurement (calculated from birth date), weight, length, head circumference, and MUAC. Nutritional biomarkers are available for some maternal and infant blood samples (see list in exploratory outcomes). Study-level variables include nutritional content of BEP, geographic setting (Africa or South-East Asia), and prevalence of undernutrition in study population (e.g., infant stunting or maternal underweight).

### IPD data collection and flow

Data for this meta-analysis will come from the four participating trials. The investigators of each trial have agreed to provide data and will send individual trial datasets to a data repository (hosted by the study sponsor). Trial investigators will remove personal identifiers, and the data repository analysts will further scan for and remove any identifiable information. Then, de-identified data will be shared with the meta-analysis research team at Penn State. After Penn State receives individual data sets, they will clean and merge them to create a pooled dataset for the IPD meta-analysis. While working with these data sets, only the Penn State IPD meta-analysis research team will have access to the individual and pooled datasets, which will be stored in a secure drive, password protected by the university.

### Data merging and quality assurance (IPD integrity)

Each trial will share their original data to the data repository team, which will prepare de-identified datasets. First, we will review the dataset processed by the data repository team to see if it includes all variables needed for the meta-analysis. Although we will receive cleaned data, we will conduct additional examination of variables quantitatively and visually by checking distributions, frequencies, missingness, and outliers (biological or statistical). For instance, for the LAZ outcome and all other z-scores, we will use the WHO criteria in determining biologically implausible values using the recommended cutoffs (LAZ <-6 or >+6; WAZ <-6 or >+5, WLZ, MUAC z-score, and head circumference z-score <-5 or >+5) (22,23). Next, we will complete initial transformations to normalize continuous distributions and categorize variables as appropriate. This step will include re-coding or creating new variables to align variable definitions for analysis with the proposed definitions from the prior harmonization work (13). Further, we will query any anomalies and compare the sample sizes and descriptive statistics with prior publications and study protocols. We will resolve any data issues or questions with the investigators for the corresponding trial. Then, we will be able to combine individual trial data sets into an analytic data set along with a variable to indicate which trial the data came from. The final product of this stage will be a merged dataset that is ready for the main analysis.

### Statistical IPD meta-analysis plan

Data validation and merging will be conducted in Stata (StataCorp, College Station, Texas); data analysis and graphic presentation of the results will be run in Stata and R (R Core Team). To investigate the effects of the BEP intervention package on outcomes, a one-stage IPD approach will be implemented for all analyses (24). This approach is preferred over the conventional two-stage IPD approach, in which trial-level aggregated summary data are pooled in the first stage for a meta-analysis model in the second stage (25,26). Overall, the use of a one-stage IPD approach will allow us to adopt more appropriate likelihood functions, make fewer model assumptions, and incorporate more effective modelling of effect modifiers to accommodate the between-study heterogeneity while we quantify the effects of BEP on maternal and child outcomes (27).

For the main analysis, the BEP intervention package will include all study arms that provided BEP to women (in the MumtaLW trial, BEP and BEP with infant azithromycin are combined; for the other trials, there is only one BEP arm). For continuous outcomes measured at six months of infant age, the effect of BEP on the outcome (e.g., LAZ) will be assessed with mixed-effects linear regression models. For dichotomous outcomes (e.g., stunting), the effect of BEP on the outcome will be assessed using relative risk estimates from mixed-effects log binomial models (or alternatively, Poisson regression if log-binomial models do not converge). We will perform both univariable analysis (BEP intervention and trial in model) and multivariable analysis (adding adjustment for baseline individual- and study-level characteristics) to examine the effect of BEP on outcomes. For multivariable models, we will include variables that are statistically significant predictors of the same outcome in the univariable analysis and variables that are unbalanced at baseline in randomized groups. In all models, intervention arm will be specified as a fixed effect, trial will be specified as a random effect (to account for heterogeneity between trials), and any other variables in multivariable models will be specified as fixed effects. Heterogeneity between trials will be assessed using *I*^*2*^ statistics. Risk of bias assessments will be carried out and reported for each of the individual trials using the Cochrane risk of bias tool.

### Subgroup Analysis: individual-level and study-level effect modification

We plan to examine effect modifiers because understanding which groups may benefit the most from BEP supplementation is an important goal of this initiative. An interaction term between the treatment and a potential effect modifier will be included in separate mixed-effects linear or log binomial (or Poisson) regression models. We will examine the following individual-level subgroups based on biological plausibility and prior literature: maternal age (<20 y, 20-29 y, ≥30 y), education (none vs ≥1 year), parity (1 child vs ≥2 children), maternal BMI (<18.5 kg/m^2^ vs ≥18.5 kg/m^2^), maternal height (<150 cm vs ≥150 cm), MUAC (<23 cm vs ≥23 cm)(28), infant sex (male vs female), and infant malnutrition as defined by low birth weight (<2,500 g vs ≥ 2,500 g), stunting (LAZ <-2 vs ≥ -2), wasting (WLZ <-2 vs ≥ -2), and underweight (WAZ <-2 vs ≥ -2). Study-level variables we plan to examine are: prevalence of infant stunting at baseline, and prevalence of maternal underweight at baseline.

In a set of exploratory analyses, we will examine if breastfeeding types (exclusive, predominant, partial) mediate the effect of BEP on the outcomes at six months. We will also analyze differences in benefits to maternal outcomes by levels of BEP adherence (<80% vs ≥80%), which can be useful in planning future studies or dissemination efforts.

### Significance level

We will set an alpha cutoff of p <0.05 to determine if our results are significantly different from those expected if the null hypothesis was correct. For testing interactions, we will use a cutoff of p <0.10. The method of Benjamini and Hochberg (29) will be applied to control for false discovery in multiple comparisons in the assessment of interaction effects in subgroup analyses.

### Missing data

The IPD meta-analysis will be analyzed based on an intent-to-treat protocol, which assumes that all randomized individuals will be included in the analyses. However, if we observe more than 20% of data missing per treatment arm, it will be flagged as potential bias (differential missingness), and we will consider two possible approaches, either performing multiple imputation assuming that data are missing-at-random (30,31), or modeling the missing data by building a Bayesian hierarchical modeling and test different missing data patterns (32). The robustness of the results will be tested in sensitivity analyses using complete cases. To estimate the missing values, the participant baseline characteristics (maternal age, maternal education, maternal BMI and height at baseline, parity, household food insecurity, baseline value of LAZ, and infant sex) will be summarized using descriptive statistics.

### Sensitivity analysis

We will test a two-stage approach in sensitivity analyses to assess the robustness of our results to different approaches. Usually, both approaches provide similar results (25,26). We will also assess the effects of variations in interventions such as BEP alone vs BEP with azithromycin vs control; BEP given with vs. without an IFA tablet; and differences in nutrient content (e.g., total calories (400 vs 600 vs 800 kcals)) in BEP products.

### Ethics and dissemination

For each individual trial, local ethical approval was obtained. This IPD meta-analysis uses de-identified data (deemed exempt from ethical approval). A centralized data repository will provide the pooled and de-identified datasets for data analysis. Further, the results of this work will seek peer-review and publication in fully open-access journals.

## DISCUSSION

Since 2016, the WHO has recommended BEP supplementation for pregnant women in settings with high rates of undernourishment to reduce incidence of small for gestational age and stillbirth. However, there are no current BEP recommendations for lactating women, and similar to pregnancy, more energy and nutrients are required in this life stage. In response, this prospective IPD meta-analysis aims to fill in that gap and investigate the effects of BEP supplementation given to lactating women. We will assess infant and maternal outcomes and identify subgroups that may benefit the most from this intervention. Prior systematic reviews and meta-analysis studies have suggested that BEP supplementation given to pregnant women may improve low birth weight, birth weight, small for gestational age, and stillbirth outcomes (8–12); improvements to maternal and child health from BEP in lactation are expected.

The four BEP trials included in our meta-analysis are conducted in low- and middle-income countries among women at risk of or with undernutrition. The BEP intervention is administered either in the form of a packaged supplement or snack, with all studies providing ≤25% of energy from protein. Unlike prior BEP research, the proposed IPD meta-analysis study has harmonized definitions for key variables of interest, and we will use concurrently conducted RCTs, which should improve the quality of data and the reliability of our estimates.

This study protocol has several strengths. Generally speaking, prospectively planned IPD meta-analysis is superior to retrospective IPD meta-analysis or aggregate meta-analysis because it is the least biased and most reliable in producing quality results. Additionally, this approach allows the combination of study- and individual-level variables from multiple trials into one dataset, thereby improving the power to assess overall effect estimates and effect modifiers. The effect modifiers can identify which groups of lactating women benefit the most from BEP supplementation (if any). We will also be able to include study-specific random effects into the analyses and investigate the influence of covariates on heterogeneity of treatment effects. These aspects will strengthen the current but limited evidence on BEP interventions and help generalize the findings in food insecure settings.

This study has a few limitations that should be considered when interpreting results. Although all studies provided BEP to lactating women, there are variations in the form and quantity of the BEP. For instance, in the IMPRINT and MumtaLW trials, BEP provides 600 and 800 kcals, respectively, which is a higher amount than the recommended range of 250-500 kcals (7). Further, the control group varies, which may impact our results. For example, IFA is not the standard of care in lactation in all settings, such as in Nepal. Also, compliance is not measured in all trials for the control group, especially when this group is advised to take advantage of the national standard of care available. Furthermore, the included trials have some differences in study design, data definitions, and data collection methods. Substantial efforts were made later to harmonize the primary and secondary outcome measures, in our case, through the BEP Initiative (13). Last, studies also differ in eligibility criteria, and one study only enrolled women that were undernourished. However, we will test in subgroup analyses if maternal undernourishment status is an important factor in the BEP treatment effect.

Ultimately, this prospective IPD meta-analysis protocol will extend our knowledge on the effectiveness of giving BEP supplementation to lactating women on infant growth and other important maternal and infant outcomes. Thus, this work aims to overcome prior challenges and clarify the benefits of BEP in dyads from low-and middle-income settings. The results of this study will be disseminated through publication in fully open-access peer-reviewed journals.

## Supporting information

PRISMA-P-checklist

## Data Availability

No data available at this stage

## Trial status

Protocol version 2. The project began on October 15, 2020. It is funded by the Bill & Melinda Gates Foundation until April 14, 2024. We preregistered the lactation meta-analysis on Open Science Framework (https://osf.io/9nq7z) on October 4, 2022. This protocol manuscript mirrors our preregistration but expands and adds important details for the included trials and data analysis methods. We have received data from three out of four participating trials via an external data repository. The data collection is on-going for the fourth trial and will conclude in the first quarter of 2024, when we anticipate to have the complete pooled dataset.

## List of abbreviations

AGP: alpha-1-acid glycoprotein
BEP: balanced energy-protein
BMGF: Bill & Melinda Gates Foundation
BMI: Body Mass Index
CRP: C-reactive protein
IFA: iron and folic acid
IPD: individual participant data
LAZ: length-for-age *z*-score
LMIC: low and middle-income countries
MUAC: mid-upper arm circumference
RCT: randomized controlled trial
WAZ: weight-for-age *z*-score
WHO: World Health Organization
WLZ: weight-for-length *z*-score

## DECLARATIONS

### Ethics approval and consent to participate

For each individual trial, local ethical approval was obtained. This IPD meta-analysis will use de-identified data (deemed exempt from ethical approval by the Institutional Review Board at Penn State). The de-identified and harmonized IPD meta-analysis dataset will be stored in a data repository. The research findings will be disseminated in peer-reviewed, open-access journals.

### Consent for publication

Not applicable

### Availability of data and materials

No data available at this stage

### Competing interests

All authors have been funded by the Bill & Melinda Gates Foundation. Parul Christian worked at the BMGF from 2015-2019 during which the BEP trials were funded. All other authors, co-authors, and collaborators have nothing to declare. JT participated on a Data Safety Monitoring Board or Advisory Board at University of North Carolina and University of California San Francisco on studies unrelated to this manuscript. JT was a non-profit board member, unpaid, for the Helen Keller International, which may use the results of these trials, depending on the results, in their programming. JT was a non-profit board member, unpaid, for the Health Volunteers Overseas that conducts work unrelated to the topic of this manuscript.

### Funding source □

This work was funded by the Bill & Melinda Gates Foundation, investment grant numberCINV-022373. The funder had no role in the study design, collection, analysis, and interpretation of data and in writing the manuscript.

### Authors’ contributors

MAC and SZ drafted the study protocol with critical guidance and input from ADG and KG. NB, PK, AM, ST, JT, RC, PC, AA, TD-C, BK, DE, FJ, JK, SK, CL, TL, MN, YS, RU contributed to the original data acquisition and to the BEP Initiative, providing input and feedback to the meta-analysis plans throughout the process. ADG led overall study design and BEP Initiative work with input from all authors. All authors read, commented, and approved the final protocol. This protocol is written on behalf of the Maternal BEP Studies Harmonization Initiative.

## Acknowledgements

We would like to thank all members of each research team for the studies included in the BEP Initiative. Our Technical Advisory Group, Drs. Martha Mwangome, Wafaie Fawzi, Sant-Rayn Pasricha, Parul Christian, and Rajiv Bahl served as scientific advisors, and we are grateful for their important guidance. Collaborators listed under **Maternal BEP Studies Harmonization Initiative:** Eleonor Zavala, Steven C. LeClerq (Johns Hopkins Bloomberg School of Public Health), Benazir Baloch (Aga Khan University), Lieven Huybregts (International Food Policy Research Institute), Laeticia C Toe (Institut de Recherche en Sciences de la Santé Burkina Faso), Giles Hanley-Cook (Ghent University), Grace J Chan (Boston Children’s Hospital), Mulatu M Derebe (Amhara Public Health Institute), Fred van Dyk, Luke C Mullany, Daniel Erchick (Johns Hopkins Bloomberg School of Public Health), Michelle S Eglovitch, Chunling Lu, Krysten North, Ingrid E Olson (Brigham and Women’s Hospital), Nebiyou Fasil, Workagenehu T Kidane, Fisseha Shiferie, Tigest Shifraw, Fitsum Tsegaye, Sitota Tsegaye (Addis Continental Institute of Public Health), Sheila Isanaka (Harvard TH Chan School of Public Health), Rose Molina, Michele Stojanov, Blair Wylie, (Beth Israel Deaconess Medical Center), Amare W Tadesse (London School of Hygiene and Tropical Medicine and Addis Continental Institute of Public Health)

## Notes

### Clinical Protocols

https://osf.io/9nq7z

### Funding Statement

This work was funded by the Bill & Melinda Gates Foundation, investment grant number INV-022373. The funder had no role in the study design, collection, analysis, and interpretation of data and in writing the manuscript.

